# Non-pharmacological interventions aimed at promoting the mental health of children and adolescents during the COVID-19 pandemic

**DOI:** 10.1101/2023.10.07.23296694

**Authors:** Carolina Villanova Quiroga, Anna Clara Sarmento Leite Caobelli, Gabriela Veiga Alano Rodrigues, Thiago Wendt Viola

## Abstract

**Background:** Interventions to promote mental health in pediatrics need to be effective, especially in crisis contexts. This systematic review proposes compile and analyze the findings of non-pharmacological interventions conducted in samples of children and adolescents during the COVID-19 pandemic, focusing on mental health.

**Method:** The research was carried out in PsycINFO, PubMed and Web of Science databases for empirical studies, including interventions in which measures of outcome variables were collected at least twice (pre and post). The studies’ samples were children and adolescents up to 19 years old, and interventions were developed throughout the COVID-19 pandemic. After eligibility analyses, 16 studies were included in this review.

**Results:** Studies used different theoretical approaches, focusing on promotion, prevention and treatment in mental health in specifics contexts. Some were delivered online, in-person, or in hybrid formats. Particularly, depression, the most frequently assessed outcome, demonstrated more favorable results within the interventions. However, due to considerable risk of bias, the analysis of results of many included studies should be performed with caution.

**Conclusions:** Most of the interventions necessitate further validation. However, the emergence of interventions during crises, such as the COVID-19 pandemic, provides an opportunity to expand evidence-based mental health practices, paving the way for their application in other crisis situations. Given that mental health prevention and promotion practices can be integrated into the roles of all healthcare providers, possessing insight into the most suitable evidence-based interventions can elevate the quality of care delivered.

## 1. BACKGROUND

During the COVID-19 pandemic, mental health has become a critical concern (Ornell et al., 2020). Abrupt changes in lifestyle, social restrictions, the fear of illness, and uncertainty about the future have created a stressful and challenging environment. A systematic review that compiled twenty-six studies measuring the effects of social distancing on the general population demonstrated an increase in symptoms of anxiety, depression, and stress resulting from preventive measures. Regarding anxiety symptoms, younger age emerged as a factor associated with higher scores, as well as student status and being female. Younger age was also associated with greater depressive and stress symptoms (Rodríguez-Fernández et al., 2021).

In this context, children and adolescents were among the age groups most affected by social isolation. They had to distance themselves from school activities, which are crucial for cognitive development, the learning process, the development of social skills, and peer interactions. Studies have shown that children and adolescents of various age groups exhibited high scores of mental health symptoms, such as anxiety, depression, and behavioral issues, during the initial phase of the pandemic (Sicouri et al., 2023). The younger the child, the higher the scores of symptoms generated by the absence of school interaction, such as anxiety (Spiteri, 2021). Younger children also struggle to verbally express their emotions, leading to more psychological and behavioral problems (Parsons, 2020). In comparison to their parents, children demonstrated higher scores of anxiety symptoms. On the other hand, adults displayed significantly higher scores of Post-Traumatic Stress Disorder (PTSD). This difference could be attributed to the cognitive skill gap between children and adults, as children’s cognitive abilities are still developing. Due to this lack of cognitive skills, children may exhibit elevated anxiety symptoms due to difficulties comprehending the issue and a lack of coping strategies (Yue et al., 2020).

In the face of emotional suffering and the potential for mental health impairment in children and adolescents, the scientific community has sought to develop and test non-pharmacological interventions (Boldt et al., 2021). These interventions aim to address a range of mental health outcomes through a multidisciplinary approach. Therefore, this systematic review aimed to compile and qualitatively assess non-pharmacological interventions developed and applied to children and adolescents during the COVID-19 pandemic. Understanding the diversity of strategies used will allow for a comprehensive evaluation of the clinical impact and effectiveness of these interventions in promoting the mental health of young individuals during times of heightened adversity.

## 2. METHODS

### Inclusion and exclusion criteria, search and screening

The PRISMA criteria for conducting systematic reviews and meta-analyses were followed, including database searches, analysis of abstracts, and selected articles. This review is registered in the PROSPERO system under registration number CRD42022356775.

The databases used in this review were PubMed, PsycINFO, and Web of Science. Empirical articles with quantitative measurement data were sought for pre- and postintervention primary and secondary outcomes. Articles with interventional methods, such as randomized clinical trials or quasi-experimental studies, were included. In this regard, theoretical reviews, cross-sectional methods, longitudinal methods, prevalence studies, observational studies, mixed methods without intervention, comparative studies, qualitative studies, case studies, editorials without original data, and intervention protocols were not considered. Additionally, data from other pandemics other than COVID-19 (e.g., 2002 SARS), articles published in languages other than English, intervention studies involving populations other than children and adolescents up to 19 years old, and intervention studies that do not focus on mental health during the COVID-19 pandemic were excluded – i.e., interventions that were already in progress or did not address the pandemic in their objectives. Furthermore, interventions that included parents and/or other family members were excluded, as well as articles that applied pre- and post-intervention measures with instruments answered by parents and/or guardians. A publication time filter was applied to the searches, restricted to works published between 2020 and 2023.

The search strategy, according to the specifics of each database, was: (‘child’ OR ‘adolescent’) AND (‘clinical trial’ OR ‘controlled clinical trial’ OR ‘randomized controlled trial’ OR ‘clinical trial protocol’ OR ‘cohort studies’ OR ‘clinical study’) AND (‘sars-cov-2’ OR ‘coronavirus’ OR ‘covid-19’) AND (‘mental health’ OR ‘psychosocial functioning’ OR ‘mental disorders’ OR ‘psychological techniques’ OR ‘psychotherapy’).

The search was conducted on June 26, 2023, by two independent judges, and is summarized in Figure 1. A third judge was invited in case of disagreements in the analyses. The abstracts were analyzed according to the pre-established inclusion and exclusion criteria, and those that met the inclusion criteria were read in full. The Rayyan QCRI program, developed by the Qatar Computing Research Institute at Hamad Bin Khalifa University, was used by the judges for reading abstracts and applying inclusion and exclusion criteria to reduce the risks of bias and analyze the material. After analysis, 26 articles were read in full. Four articles identified in reference lists were manually included for a full reading. Of these, 14 were excluded, totaling 16 studies included in the results of this review.

**Figure 1.**
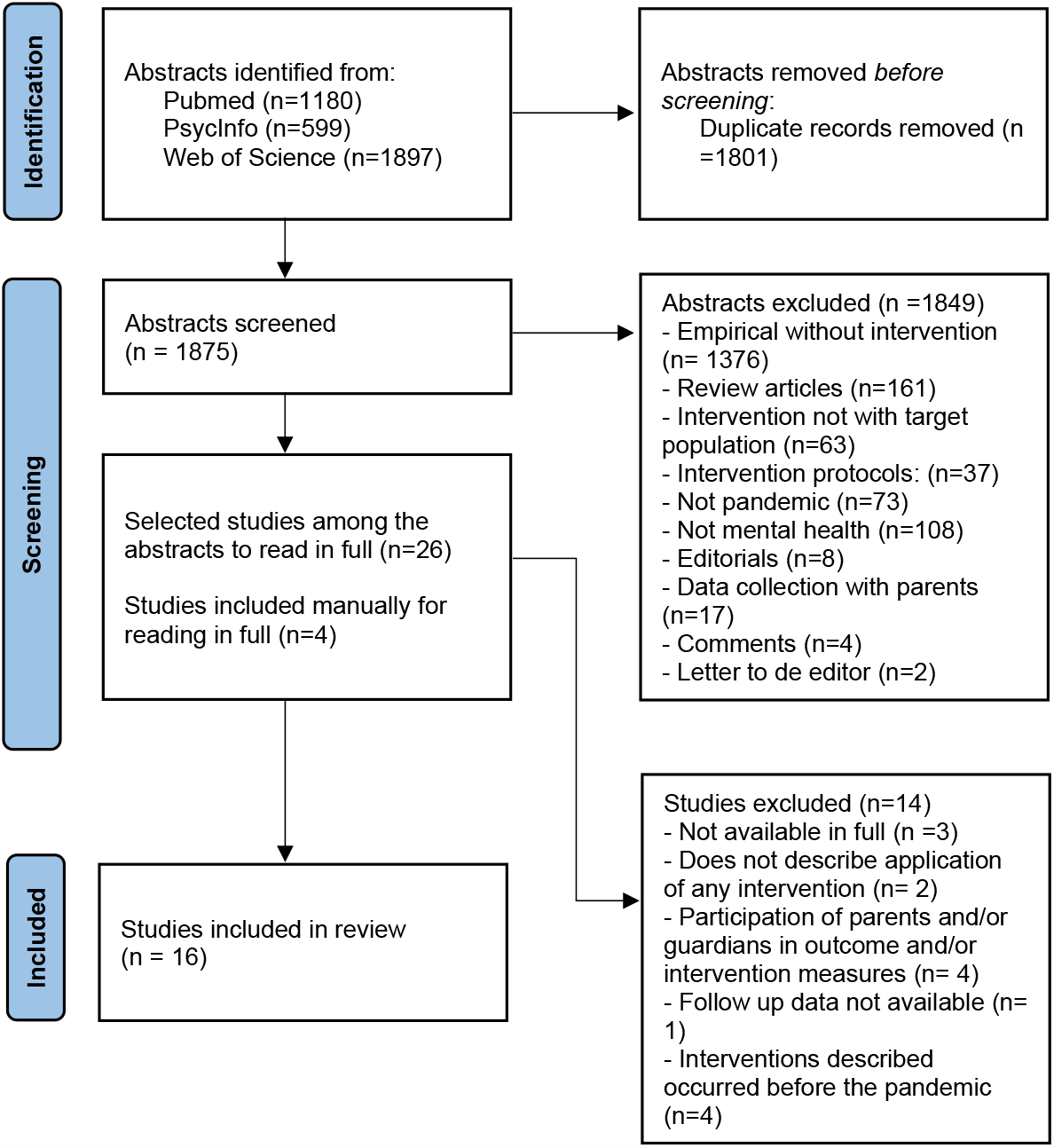
PRISMA-based flowchart.

The Revised Cochrane risk-of-bias tool for randomized trials (RoB 2) – version 9 (Sterne et al., 2019) was used to assess the risk of bias in the studies. The tool consists of five domains: risk of bias arising from the randomization process, risk of bias due to deviations from the intended interventions (effect of assignment to intervention), missing outcome data, risk of bias in the measurement of the outcome, and risk of bias in the selection of the reported result. In the end, an overall risk of bias is determined. Each domain contains different questions that can be answered with yes, probably yes, probably no, no, or no information.

This tool is intended for the assessment of randomized studies. However, as this review also included single-group studies that provided follow-up measurements, the tool was adapted – in these cases, domain one, concerning the randomization process, was left blank.

There is also a cluster-randomized study included in the review. Cochrane recommends another tool for evaluating this specific method – however, since it involves only one study, RoB 2 was also adapted. Furthermore, it was assumed that the objective of the review was to assess the effect of assignment to intervention (the ‘intention-to-treat’ effect) in all included studies. Finally, in the analysis of all studies, it was noted that the only source of information to help inform bias risk was the journal article – randomized clinical trial protocols were accessed only in studies that reported their number.

### Analysis

The analysis of the selected articles was conducted qualitatively. These included the country of publication, methods, composition of the sample, definition of outcome variables, specific details about the proposed interventions, and the outcomes they yielded. When considering the composition of the sample, factors such as its clinical or non-clinical nature, the inclusion of specific genders or both, age range, and sample size, were registered. Regarding the intervention, details about the duration, frequency, setting, and goals, were registered. In terms of the outcomes, information about whether the intervention was successful in reducing psychological symptoms like depression or anxiety, were included in our data extraction protocol.

## 3. RESULTS

### 3.1 Methodological Quality and Risk of Bias

Table 1 presents the results of the risk of bias assessment for the included studies. As indicated, no study demonstrated a low risk of bias. Six studies exhibited some concerns, while ten showed a high bias risk. Non-randomized studies presented more significant risks. However, several randomized trials did not adequately describe the randomization process or study protocols. Moreover, few studies reported pre-defined outcome analysis plans, leading to a risk of data bias in nearly all analyzed works.

**Table 1.**
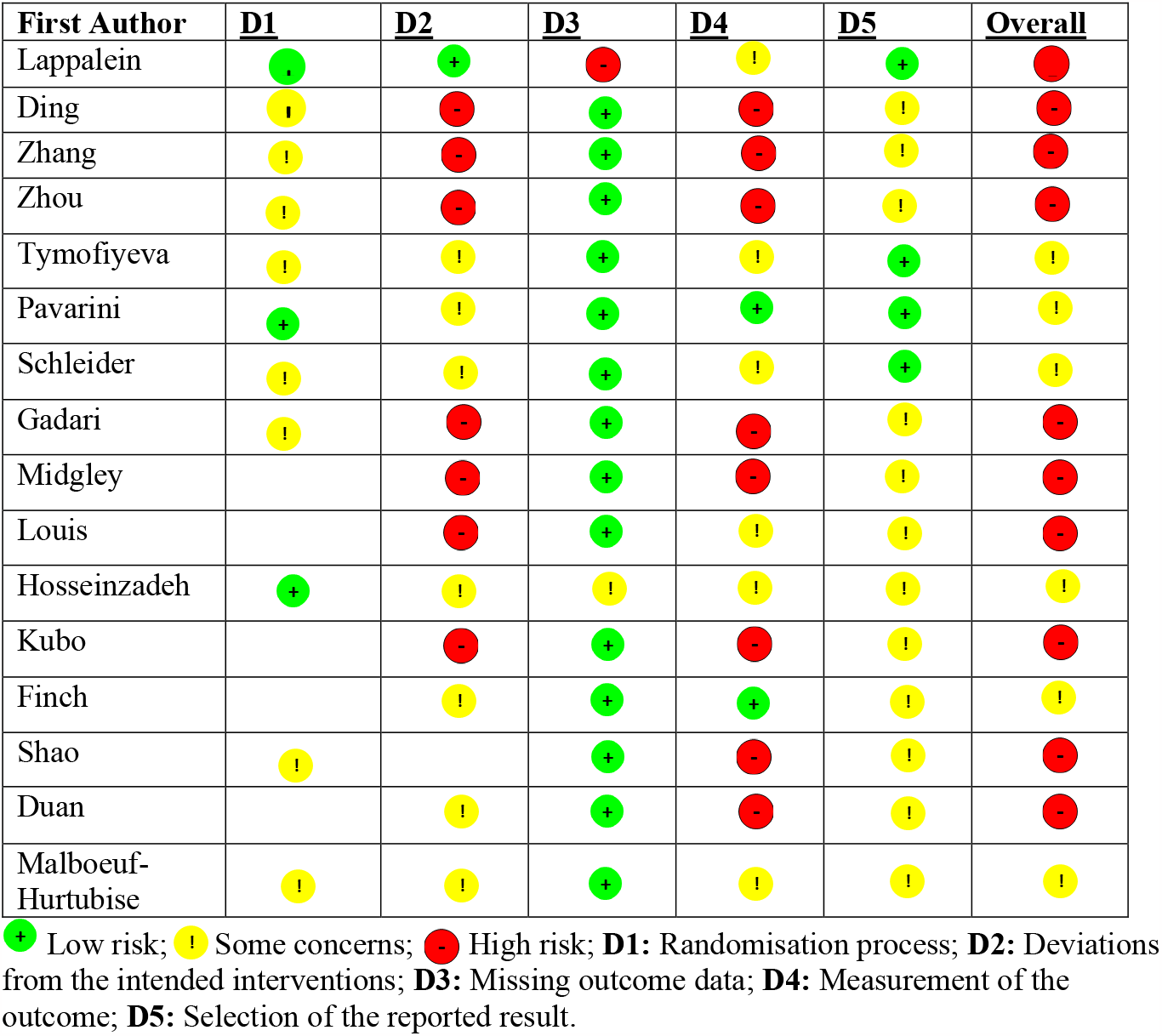
Risk of bias and methodology quality of the selected studies.

### 3.2 Studies and Interventions Characterization

Table 2 provides general information about the studies included in this review. Although all interventions took place during the COVID-19 pandemic, some were delivered online, in-person, or in hybrid formats. Notably, a wide range of theoretical frameworks underpinned the proposed interventions.

**Table 2.** Data from the included studies.

| **First Author/Year** | **Study design** | **Country** | **Objective** | **Population** | **Intervention** | **Outcome variables** | **Main results** |
| --- | --- | --- | --- | --- | --- | --- | --- |
| Lappalainen (2023) | Randomized clinical trial | Finland | Investigate the effects of an online ACT intervention to promote psychological flexibility and self-compassion in adolescents and reduce psychological distress during the second wave of COVID-19. | Adolescents aged between 14-16 years old attending the eighth grade. N=234. | Three groups: 1) a 5-week Youth Compass online intervention with two 45-minute video calls from a learner coach and virtual coach support (iACT learner coach + virtual coach group); 2) a five-week Youth Compass online intervention with a 15-minute video call from a student coach and support from a virtual coach (iACT virtual coach group); and 3) a group without intervention | Anxiety, depressive symptoms, psychological flexibility, and self-compassion | In isolation, groups 1 and 2 did not differ from the control group in any measured variable. However, when combining groups 1 and 2 and looking at teens who completed at least 30% of the Youth Compass, there were changes in anxiety scores, greater psychological flexibility, and self-compassion. |
| Ding (2020) | Randomized clinical trial | China | Provide referrals to improve adolescents' mental health levels during a crisis event. | Adolescents between 12-18 years old, with scores for anxiety symptoms. N= 150 | The clinical group participated in an eight-week peer education and aerobic exercise intervention. The control group received routine health education. | Anxiety, depression, and sleep quality. | There was an improvement in anxiety and depression scores in both groups, but with a more significant difference in the clinical group. The clinical group also showed better results in terms of sleep quality. |
| Zhang (2021) | Randomized clinical trial | China | Analyze the effect of research-based psychological counseling | Adolescents between 12-18 years old with symptoms of | The control group received routine community health education. The clinical group received an eight- | Resilience, depression, anxiety, and sleep quality. | The clinical group showed improvement in depressive and anxiety scores compared to the control group, as well as sleep |
|  |  |  | intervention on adolescent mental health during the COVID-19 pandemic. | anxiety. N=160 | week intervention, with psychological training in a research-based psychological counseling model and outdoor exercises. |  | quality and psychological resilience. |
| Zhou (2022) | Randomized clinical trial | China | To explore whether mindfulness intervention is an efficient method to increase emotional intelligence and psychological capital in adolescents. | All eighth and ninth-grade students in a school, ages 14-15. N=798. | The clinical group received mindfulness lessons twice a week in face-to-face psychology classes. The control group received no intervention. | Emotional intelligence and psychological capital | There was an increase in emotional intelligence scores at all times in the intervention group and psychological capital. The control group showed no improvement over time. |
| Tymofiyeva (2022) | Randomized waitlist-controlled trial | United States | (1) to test the feasibility of TARA, delivered partially over Zoom, and (2) to assess changes in the emotional well-being of healthy adolescents between the ages of 14–18 during the COVID-19 pandemic. | Healthy adolescents aged 14-18 years old. N=21 | The intervention group received a mindfulness intervention based on the neuroscience TARA. The intervention was hybrid due to the pandemic. The control group was on a waiting list. | Emotional wellbeing | No significant differences were found in measures of well-being between the intervention group and the passive control group. However, improvement in functioning was measured within the intervention group. |
| Pavarini (2023) | Randomized clinical trial | United Kingdom | Test the short-term effectiveness of an online training program to equip young people with skills to support their peers' mental well-being during the COVID-19 pandemic. | Adolescents aged 16-18 years old from anywhere in the UK. N=100. | Intervention group: interactive sessions for all participants with information + smaller groups for practical and sharing activities. Waiting list control group. | Primary outcome: motivation to provide support, perceived ability to provide support, frequency of support provided, compassion for others, and connection with peers. Secondary outcome: mental well-being, emotional symptoms, self-efficacy, and civic engagement | The training effectively increased the youth's ability to support peers during the COVID-19 pandemic. They felt more confident in their supportive abilities, showed greater compassion towards others, and felt more connected with their peers. Training also showed a positive effect in improving mental well-being and reducing negative emotional symptoms. Furthermore, participants in the training group reported greater self-efficacy and civic engagement than the control group. |
| Schleider (2022) | Randomized controlled trial | United States | Test single-session online interventions during COVID-19 in adolescents with elevated symptoms of depression. | Adolescents aged 13-16 years old with high depressive symptoms. N=2452. | Group 1: SSI-BA that addresses activity withdrawal and low agency; Group 2: SSI-GM designed to address hopelessness; Group 3: control. | Depression, hopelessness, anxiety, trauma related to covid, and restrictive eating. | The SSI-BA and SSI-GM interventions effectively reduced depressive symptoms compared with the placebo intervention. There were no significant differences between the two interventions regarding these outcomes. Furthermore, the beneficial effects of the primary outcome measures were maintained at the three-month follow-up after the intervention. |
| Gadari (2022) | Randomized experimental study | Iran | To determine the effect of virtual resilience training on the social self-efficacy of elementary school girls. | Girls aged 9-10 years old, students at a school chosen for convenience. N=80. | Twelve virtual resilience training sessions over 6 weeks, twice a week. The training was delivered through video and audio clips, animations, children's scenarios, text messages and storytelling. The Control group had no intervention. | Social self-efficacy | Social self-efficacy significantly increased after the intervention. The control group showed no significant difference. |
| Midgley (2021) | Single-group, uncontrolled exploratory design | United Kingdom | To examine the feasibility, acceptability, and effectiveness of an English-language adaptation of iPDT for depressed adolescents. | Adolescents between the ages 16-18 who met the criteria for Major Depressive Disorder. N=23. | The online intervention includes videos and texts on a specific topic, complemented by worksheets that young people fill out and send to their therapeutic support agent, who provides feedback. In addition, participants have a weekly 30-minute text "chat session" with their therapeutic support worker. | Primary outcome: depression; Secondary outcomes: anxiety and emotional regulation. | Improvement in depression scores and emotion regulation, but no significant differences in anxiety scores. |
| Louis (2023) | Single-Group Pilot Study with Pre- and Post-Test Measures | India | Develop and pilot test an online intervention program to increase the self- | Adolescents aged 11-17 were chosen from six charitable shelters in | Online cognitive self-compassion intervention program. | Self-esteem | There was an increase in self-esteem scores in the pre-test and post-test of participants. |
|  |  |  | esteem of adolescents exposed to parental intimate partner violence. | Kerala, India, exposed to parental intimate partner violence. N=10. |  |  |  |
| Hosseinzadeh (2023) | Single-blind randomized clinical trial | Iran | Investigate the effects of virtual logotherapy on the health-promoting lifestyle of girls raised by single parents during the COVID-19 pandemic. | Girls aged 13-18 years old who have only one parent. N=88. | The clinical group received online logotherapy interventions in groups of 3-5 people. | Health-promoting lifestyle | After the intervention, the health-promoting lifestyle scores and all the dimensions that comprise the assessment instrument were significantly higher in the intervention group. |
| Kubo (2022) | Intervention study | Japan | Investigate whether a single school-based intervention, including self-monitoring and psychoeducation for COVID-19, effectively promoted children's mental health. | All students from 1st to 3rd grades of a school, aged 12-15 years old. | 3rd series: a 15-minute self-monitoring session and a 35-minute psychoeducation session; 2nd grade: received a 15-minute announcement stating that pandemic-related stress affects mental health; 1st series: control group without intervention. | Depression, fear related to covid, and anxiety. | The intervention group significantly reduced depressive symptoms between t1 and t2. There was no significant effect on fear of COVID-19 and trait anxiety. |
| Finch (2023) | Non-randomized pre- and post-test design | Australia | To examine the effectiveness of an intervention aimed at PsyCap and assess its impact on | Girls from a Catholic school. N=131. | MY HERO intervention program consists of four workshops in the classroom, lasting 50 minutes each, namely: workshop on hope, | Primary outcome: anxiety, depression, and subjective | There were no significant changes in the primary outcome variables. There was an increase in optimism and efficacy and a decrease in self-oriented perfectionism. Hope |
|  |  |  | mental health symptoms and subjective well-being in a class of senior high school students. |  | efficacy, resilience, and optimism. All workshops included a guided worksheet activity for students, aiming to develop the session construct using Cognitive Behavioral Therapy techniques. | well-being. Secondary outcome: hope, effectiveness, resilience, optimism, and perfectionism | approached statistical significance. |
| Shao (2021) | Randomized controlled trial | China | To evaluate the effect of a dance intervention based on the Satir Model on the mental health of adolescents with depression during the COVID-19 pandemic. | Adolescents with depressive symptoms. N=62. | The experimental group received intervention combining group psychological therapy and dance therapy. | Depression, anxiety, mental health, life satisfaction, and psychological resilience. | After the intervention, the experimental group's scores on anxiety and depression were lower compared to the control group. The experimental group's scores on life satisfaction, psychological resilience, and its dimensions are higher than those of the control group. |
| Duan (2022) | A quasi-experimental study with no control group | China | To develop and investigate changes in anxiety symptoms and quality of life among participants in force-informed ACT in an online program. | 5th-grade primary school students aged 10-12 years old. N=76. | The online intervention combines elements of ACT theory with a strengths-based approach. The process involves two main aspects: (a) Mindfulness and Acceptance Processes and (b) Commitment and Behavior Change Processes. Six main techniques are used, which aim to achieve cognitive and behavioral changes. | Anxiety and quality of life | Significant reduction in anxiety symptoms but not a statistically significant increase in quality of life. |
| Malboeuf-Hurtubise (2021) | Randomized cluster trial | Canada | Analyze the effects of P4C and MBI online interventions on mental health in the context of the COVID-19 pandemic. | Elementary school students from two schools. N=37. | Online interventions based on MB and P4C were applied. Both had 5 weekly sessions of 1 hour, online and in real-time. | Inattention, anxiety, and psychological need satisfaction | The P4C group significantly decreased mental health difficulties, while the MBI group showed no significant changes. The MBI group reported greater satisfaction with basic psychological needs after the intervention, while the P4C group did not observe significant changes in this construct. |

ACT: Acceptance and Commitment Therapy; iACT: online-delivered ACT intervention; iPDT: internet-based psychodynamic treatment; MBI: Mindfulness-Based Intervention; PsyCap: Psychological Capital; P4C: Philosophy for Children; SSI-BA: Single Session Behavioral Activation Intervention; SSI-GM: Mental Growth Single Session Intervention; TARA: Training for Awareness, Resilience, and Action.

One study implemented an intervention based on a humanistic theory protocol (Shao, 2021); another was grounded in psychodynamic theory (Midgley et al., 2021); logotherapy was used in one study (Hosseinzadeh et al., 2023); two studies employed acceptance and commitment therapy (ACT) (Duan et al., 2022; Lappalainen et al., 2023); self-compassion was the focus of one study (Louis & Reyes, 2023); and three studies utilized mindfulness as the foundational premise of the interventions (Malboeuf-Hurtubise et al., 2021; Tymofiyeva et al., 2022; Zhou et al., 2022). Additionally, seven studies (Ding & Yao, 2020; Finch et al., 2023; Gadari et al., 2022; Kubo et al., 2022; Pavarini et al., 2023; Schleider et al., 2022; Zhang et al., 2021) did not specify a foundational theory but relied on assumptions like behavioral activation and psychoeducation. Notably, no studies indicated cognitive-behavioral therapy (CBT) as the theoretical basis for constructing or developing the proposed intervention. Only one study (Finch et al., 2023) mentioned that the techniques employed were derived from CBT.

Three studies (Ding & Yao, 2020; Shao, 2021; Zhang et al., 2021) featured interventions combining mental health professional-delivered protocols with physical exercises. For instance, Shao described a humanistic theory-based intervention alongside dance activities. The author argued that dance practice allows non-verbal expression for adolescents who struggle with communication, facilitating engagement, and enhancing outcomes.

Four studies aimed to intervene with clinical samples, including three targeting adolescents screening for depressive symptoms (Midgley et al., 2021; Schleider et al., 2022; Shao, 2021) and one focusing on adolescents with anxiety symptoms (Zhang et al., 2021). It is worth noting that three studies (Finch et al., 2023; Gadari et al., 2022; Hosseinzadeh et al., 2023) exclusively selected samples of girls. Among these, only Hosseinzadeh et al. specifically sought girls with only one parent. The authors believed girls were more susceptible to emotional distress and mental health symptoms during crises such as the COVID-19 pandemic. Although most selected studies reported the percentage of boys and girls in their samples, few performed statistical analyses comparing gender effects, with Kubo et al. (2022) being an exception. They reported no significant gender differences in baseline measures of depression, anxiety, and COVID-19-related fear.

Amid the pandemic context, many studies tested data collection and intervention, which were presented online or in hybrid formats. However, some studies in this review needed more precise descriptions of these details. Given the potential for both in-person and online applications, as exemplified in the work of Tymofiyeva et al. (2022), an accurate description of how the intervention was performed in this matter is essential. Two studies that combined psychological interventions with physical exercises (Kubo et al., 2022; Shao, 2021) did not clarify whether professionals delivering the modalities were in-person with study participants, nor whether instruments used to measure outcome variables were administered in-person or online. Other studies described intervention delivery but did not detail outcome evaluation instruments (Duan et al., 2022; Hosseinzadeh et al., 2023).

### 3.3 Outcomes and Findings of Interventions

Five studies (Gadari et al., 2022; Hosseinzadeh et al., 2023; Louis & Reyes, 2023; Tymofiyeva et al., 2022; Zhou et al., 2022) employed positive aspects of mental health promotion as outcome variables rather than evaluating psychopathological symptoms as primary or secondary outcomes. These positive outcomes included quality of life, self-esteem, subjective well-being, self-efficacy, compassion, life satisfaction, and emotional intelligence. Other studies assessed outcomes related to the severity of depression symptoms, anxiety, sleep quality, inattention, hopelessness, restrictive eating, and COVID-19-related fear.

Among studies that used depression as an outcome variable, six (Ding & Yao, 2020; Kubo et al., 2022; Midgley et al., 2021; Schleider et al., 2022; Shao, 2021; Zhang et al., 2021) reported positive effects in reducing symptoms, making it the most promising psychopathological variable. However, one study (Finch et al., 2020) reported no effects of the tested intervention on reducing depressive symptoms. Studies evaluating improvements in sleep quality (Ding & Yao, 2020; Zhang et al., 2021) and restrictive eating (Schleider et al., 2022) yielded positive results.

Regarding anxiety as an outcome, the results were less promising. Five (Ding & Yao, 2020; Duan et al., 2022; Lappalainen et al., 2023; Shao, 2021; Zhang et al., 2021) studies reported improved symptomatology, while three (Finch et al., 2023; Kubo et al., 2022; Midgley et al., 2021) did not achieve positive outcomes. Schleider et al. (2022) comparing two different interventions with a control group, obtained varied anxiety results. One group demonstrated improvement, while the other did not. A similar pattern emerged for COVID-19-related fear, which did not improve. This variable also failed to show positive results in Kubo et al (2022).

Studies that adopted positive mental health-related aspects as primary or secondary variables yielded significant insights into the outcomes of the tested interventions. Tymofiyeva et al. (2022) were the only ones to adopt well-being as a sole outcome variable but found no significant differences between the experiential and control groups post-intervention. However, in the study by Pavarini et al. (2023), where well-being was a secondary outcome, positive results were observed. Duan et al. (2022) chose quality of life as the primary outcome and anxiety as a secondary outcome. Their intervention effectively reduced psychopathological symptoms but did not yield significant results in the primary outcome variable.

Kubo et al. (2022) and Schleider et al. (2022) were the sole studies to test the effects of a single-session intervention, albeit one in-person and the other online. The former aimed to ascertain whether a single session could promote mental health and reduce COVID-19-related fear in school-aged children. The authors used depression, anxiety, and COVID-19-related fear symptoms as outcome measures. The latter aimed to test the efficacy of an online intervention focused on teaching a growth mindset to adolescents with depression symptoms.

## 4. DISCUSSION

This systematic review aimed to compile and qualitatively evaluate non-pharmacological interventions developed and applied to children and adolescents during the COVID-19 pandemic. After database searches and applying inclusion and exclusion criteria, 16 studies were included and analyzed for bias risk, methodological quality, and details of proposed interventions. Overall, both studies that focused on psychopathological symptoms as outcomes and those that examined positive aspects of mental health showed promising results. Particularly, depression, the most frequently assessed outcome, demonstrated more favorable results.

Attention is drawn to the low methodological quality of the included studies, as well as the high risk of bias. With the inclusion of non-randomized single-group studies, it was hypothesized that these would present more significant bias in their results. Even randomized studies displayed substantial biases, which should be considered when assessing the effectiveness of the interventions in question. Some studies claimed to be randomized but did not describe the randomization process. Others failed to address the impact of sample loss throughout the process. Given the pandemic circumstances and challenges in conducting research as usual, some fundamental processes might have been compromised, affecting methodological quality. Consequently, the results reported in these studies should be interpreted with caution.

The assessment of bias risk is crucial for validating the efficacy of an intervention. How an intervention is conducted, the potential for blinding to minimize interference, the publication of a protocol detailing randomization and intended statistical analyses, and the evaluation of sample loss and its impact on results determine the effectiveness of the tested model and its potential for replication.

While Cognitive Behavioral Therapy (CBT) has a significant body of research with proven efficacy in interventions with children and adolescents (Bastien et al., 2020), this systematic review included articles describing interventions based on different theoretical frameworks, such as psychodynamic, humanistic, and Acceptance and Commitment Therapy (ACT). Only one study mentioned the use of CBT techniques (Finch et al., 2023), but none described CBT as the theoretical foundation for the developed interventions.

The possibility to test interventions, particularly in a crisis context like the COVID-19 pandemic, is crucial to address potential long-term impacts that are not yet understood. There is no similar recent historical precedent to provide precise theoretical and technical support for the mental health effects on the population. Additionally, children and adolescents are a particularly vulnerable population with potential psychosocial repercussions (Stavridou et al., 2020). Therefore, studies that address mental health promotion and prevention are especially important. However, it is essential to prioritize evidence-based practice when professionals apply specific techniques in their clinical work, even in crises, to ensure the selection of the best evidence and consider the population’s characteristics and preferences.

Especially in the field of mental health, the American Psychological Association (APA) established the Presidential Task Force on Evidence-Based Practice (APA, 2006) in 2005, defining evidence-based practice as “the integration of the best available research with clinical expertise in the context of patient characteristics, culture, and preferences.” Attention to evidence-based practice helps assess the use of interventions and the possibility of extending their use to other samples. Considering the social and health context in which the interventions in this review occurred, it is important to discuss their long-term applicability. Promoting mental health during the COVID-19 pandemic is crucial, but the ability to replicate these interventions outside of crisis situations amplifies their impact.

We also highlight the need for more studies with group therapies, as they raise important issues such as ethics, confidentiality, and therapeutic alliance (Weinberg, 2020). A metaanalysis study indicated that concerning positive psychology interventions, individuals engaged in long-duration therapeutic processes, whether individual or group-based, show more promising results (Carr et al., 2021). This systematic review identified studies with different formats and various primary and secondary outcome variables. The ability to assess the efficacy of group or individual interventions, whether short or long-term, highlights the range of possibilities for mental health intervention and contributes to the development of efficacy research.

However, perhaps the most interesting outcome is evaluating the potential for effective delivery of mental health interventions remotely. Due to the need for social isolation, many studies and mental health interventions were conducted online. In this review, most studies described interventions through digital platforms such as Zoom or WhatsApp. In times of pandemic, providing mental health assistance online allows access to services and mitigates negative effects.

Before COVID-19, previous studies on children and adolescents found that online interventions were well-accepted and yielded promising results in terms of positive mental health outcomes (Grové & Reupert, 2017; Sweeney et al., 2019). Indeed, this system is adaptable, quick, and cost-effective (Langarizadeh et al., 2017). However, the delivery of online mental health interventions during the pandemic raises interesting discussions, especially given the growing investment in this model. Authors argue that the chaotic and crowded home environment during social isolation may have influenced session dynamics (Perrin et al., 2020). Other challenges are related to technology itself, as the effectiveness of online interventions also depends on access to technological devices like computers and smartphones, as well as quality internet connections that facilitate engagement in treatment. In the context of the COVID-19 pandemic, access to these resources or lack thereof highlights social disparities that need to be considered in research, as they directly impact sample heterogeneity. However, overall, the pandemic has accelerated the adoption of digital strategies in mental health, prompting technical developments that warrant further attention and refinement.

*Limitations and Strengths*

This study presents limitations that deserve attention. Only three databases were used, potentially excluding interventions not indexed in those databases that could have been included in this review. Studies in which parents and/or guardians participated in data collection and/or intervention were excluded, possibly omitting promising research that could have contributed to the discussion. Furthermore, the incorporation of studies with diverse methodological designs renders statistical tests like meta-analysis unfeasible for measuring the effect size of the included interventions. Despite the limitations, this review was conducted with methodological rigor, following the PRISMA criteria and utilizing the Cochrane tool for bias assessment. In addition, the protocol was previously registered with PROSPERO.

## 5. CONCLUSION

The COVID-19 pandemic posed an intense challenge in the health and mental health field due to its unpredictability, high mortality rates, and difficulty in delivering routine healthcare services. The necessity of maintaining social isolation measures for contagion prevention resulted in significant mental health repercussions, driving exploratory and intervention research efforts to understand and address them. Particularly, children and adolescents proved to be a vulnerable population, experiencing significant psychological and social effects (Stavridou et al., 2020).

Developing new interventions focusing on mental health prevention and promotion is crucial in this context. However, several important considerations emerge in this systematic review that aimed to compile and evaluate non-pharmacological interventions for the mental health of school-age children and adolescents. Given the overall high risk of bias in the presented interventions, the results should be interpreted cautiously. Future studies, with a focus on long-term repercussions, are essential. Furthermore, replicating the analyzed interventions in different samples and contexts could contribute to new data that may support their significance in addressing the mental health of children and adolescents during moments of crisis.

### Financing

This research did not receive any specific funding from funding agencies in the public, commercial, or not-for-profit sectors.

### Key Messages

- The COVID-19 pandemic has been emotionally demanding for children and adolescents.
- Social isolation caused different damages to mental health.
- Non-pharmacological interventions are an effective form of health promotion.
- Interventions that promote mental health in pediatrics should be evidence-based.
- Interventions developed during COVID-19 can be applied in other crisis situations.

## Data Availability

All data produced in the present work are contained in the manuscript

